# Why do border controls fail to stabilise epidemics?

**DOI:** 10.1101/2025.05.12.25327440

**Authors:** Kris V Parag, Patrick Hoscheit

## Abstract

Can border controls and travel restrictions suppress epidemics? This is an important and recurring question for public health policymaking and pandemic preparedness. Although there is a consensus that infections imported from external locations are critical to sustaining local epidemic growth and driving the endemic equilibria of the disease, there remains ongoing debate on the effectiveness of interventions aimed at curbing importations. Most studies contributing to this debate rely on complex metapopulation models that preclude generalisable insights and formal control principles are rarely used. Here we demonstrate how classical control theory applied to an analytic but flexible and widely used transmission model provides conclusive evidence that border and travel restrictions fail to stabilise spread. These restrictions always enter as precompensators and hence fail to shift the epidemic poles. Consequently, regions cannot respond to emergent outbreaks in isolation. We find that coordinating interventions across both the local and external regions converts precompensation into feedback control and derive new criteria for overall stability. We further develop formulae specifying how border controls shape performance, such as disease equilibria and total infections. During growing epidemic phases, cooperative control is crucial for stability. Once stability is assured, border control can be sufficient to meet performance targets.

## I. Introduction

Although border controls, screening and travel restrictions are among the most widely implemented measures in response to outbreaks of directly transmitted infectious diseases, their effectiveness at controlling spread remains an active topic of debate [1]–[3]. During the growing and decaying phases of an epidemic, infections introduced from external regions can spur and sustain local transmission [4]. Repeated importations can drive epidemic waves in previously unaffected locations, precipitate pandemics and cause seasonal cycles among source and sink locations. While the use case for restricting mobility is clear, opinion is divided on the usefulness of these measures.

Some studies have found that even stringent border controls are ineffective against transmission and only delay epidemics by a few weeks [1], [3], [5]. Other works have linked timely travel restrictions to reduced spread [2], [6], [7], building a case for border control as a central part of epidemic response strategy. A third strand of research has questioned whether past data have been of sufficient fidelity to meaningfully infer and extricate the impact of border control from local interventions, highlighting the value of enhanced import surveillance [3], [4]. However, there is a consensus that epidemiological modelling frameworks are a key tool for resolving this debate (even as data improve) and informing border control policy [5], [6].

The most common modelling approaches employ complex metapopulation or agent-based models [5], [8]. While valuable for detailed simulation of disease transmission, these models can be hard to validate and unsuitable for extracting generalisable insights. Here we aim to uncover the fundamental limits on what border and travel controls can and cannot achieve by applying classical control theory. Although control theory has found success in epidemiology, it has been underutilised in this field [9], [10]. By deriving new epidemic transfer functions and identifying stability and performance thresholds, we show how standard control analyses provide a definitive and testable answer about the effectiveness of border closures.

## II. Model Formulation

### A. Uncontrolled epidemic dynamics

We describe the transmission dynamics of a generic infectious disease using the renewal model [11]. This is a discrete-time autoregressive process in which new local infections at time *t, i*(*t*), result from imported infections *m*(*t*) and the convolution of past infections with a kernel *w*(*t*) as in (1).

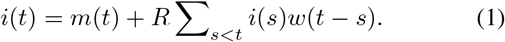

The kernel determines the distribution of the times between primary and secondary infections i.e., *w*(*t* − *s*) is the probability that it takes *t* − *s* time units for an infection to be transmitted. The parameter *R* is the reproduction number of the disease and defines the level of branching in transmission. A single infection generates *R*Σ_*s*≤*d*_ *w*(*s*) = *R* new infections across the period *d* that it is infectious (after which recovery occurs and the individual is no longer infectious).

The renewal model assumes that infectious and susceptible individuals mix randomly and generalises many popular disease models (e.g., the susceptible-exposed-infected-recovered (SEIR) model) [12]. The dynamics of COVID-19, Ebola virus disease, SARS, measles and HIV, among others, have all been simulated by appropriately specifying *R* and the generation time distribution *w*(*t*) [13]. We focus on initial epidemic stages so that *R* is fixed until we introduce control (i.e., the fraction of individuals infected is small relative to the total population size). We can use piecewise-constant switches in *R* to approximate later epidemic stages and this system is often used to determine asymptotic epidemic properties [14].

We construct the epidemic transfer function *G*(*z*) [15] by taking the Z transform of (1) with capitalised forms denoting the transformation e.g., 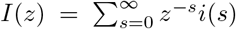. This yields (2), which has *W* (1) = 1 because it is the Z transform of a distribution. We also know that *RW* (*r*) = 1 from the Euler-Lotka relation [16] with *r* as the dominant pole of *G*(*z*).

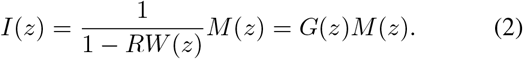

This real pole is the multiplicative (asymptotic) growth rate *r* of the disease (this is the discrete time form of the commonly used growth rate [17]). Critical stability therefore occurs when *z* = *r* = 1 i.e., the dominant pole sits on the unit circle. This corresponds to *R* = 1 because *RW* (1) = 1. The system is asymptotically stable at *r <* 1 or equivalently *R <* 1.

The relative stability of (2) is set by how close |− *RW* (*z*)| is to −1 and this system has a phase crossover frequency of 0 [15] so *R*^−1^*W* (*e*^0*j*^)^−1^ = *R*^−1^ (the gain margin). This is the control effort required to force the epidemic to critical stability and is realisable by removing a proportion 1 − *R*^−1^ of new infections over the infectious period *d*. This fraction relates to herd immunity and other epidemic control thresholds and leads to a controlled growth rate of *r*_*c*_ = 1. The controlled reproduction number *R*_*c*_ is the inverse of how much we can scale infections to force critical stability and also equals 1 [15]. The delay margin of (2) (when stable) is infinite. We generalise this model in the next sections to analyse interventions and derive new stability and performance criteria.

### B. Interventions on local and imported cases

The model above is idealistic because (i) we rarely observe actual infections, (ii) we have not considered how interventions or control actions alter transmission (to obtain *R*_*c*_) and (iii) we have ignored how infections imported into the location of interest can influence interventions. Here we incorporate (i)-(iii) to derive a more realistic and general controlled renewal model. A key novelty of this model is that we distinguish between control actions that limit the infections *m*(*t*) imported from source regions and controls that suppress the net infections *e*(*t*) = *i*(*t*) − *m*(*t*) generated in our local or sink region.

The most commonly recorded signal during an unfolding epidemic is the time series of symptomatic cases. Cases are under-reported and delayed indicators of infections [18], [19]. We let *h*_*e*_(*x* − *s*) be the cumulative probability that locally generated infections at time *s* are detected as cases by time *x* and define *c*_*e*_(*x*) = *e*(*s*)*h*_*e*_(*x* − *s*). We describe a wide array of detection schemes as 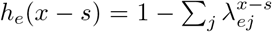, with the *λ*_*ej*_ as detection rates. This is the cumulative probability of detection under an Erlang distribution, the simplest of which is a geometric distribution with 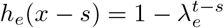.

We allow for differences in the detection of imported infections as cases because border screening or testing, for example, may be different in effectiveness from local case reporting or contact tracing. Hence, we also define *c*_*m*_(*x*) = *m*(*s*)*h*_*m*_(*x* − *s*) with the geometric detection scheme 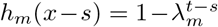. As (*λ*_*e*_, *λ*_*m*_) → (0, 0), (*c*_*e*_(*x*), *c*_*m*_(*x*)) → (*e*(*x*), *m*(*x*)) and detection is perfect. At the other extreme, (*λ*_*e*_, *λ*_*m*_) → (1, 1) and no infections are detected. For ease of notation we write all subsequent results for the geometric distribution but note that extension to Erlang and more general discrete time phasetype distributions all follow directly and easily.

Detection is important because targeted interventions such as testing, tracing and isolation, pharmaceutical administration or quarantines only apply to observable infections. We model generic interventions as a removal of the proportions *u*_*e*_ and *u*_*m*_ of locally generated and imported cases, respectively. As a result, infections *i*(*s*) are subject to control actions of effect −*u*_*e*_*h*_*e*_(*t* − *s*)*e*(*s*) −*u*_*m*_*h*_*m*_(*t* − *s*)*m*(*s*). We therefore obtain the controlled renewal model of (3), which generalises (1).

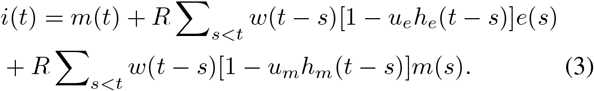

Maximum control effort applies *u*_*e*_ = *u*_*m*_ = 1 and minimum occurs when both are 0 (and recovers (1)). As far as we can tell, (3) is novel and expands on frameworks from [15], [20]. Unobserved local or imported infections transmit without any reduction as they are not identified as cases by targeted control schemes. Applying Z transforms to (3) and observing that the cumulative detection probabilities result in a time dilation of the generation time distribution we derive (4).

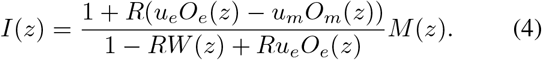

We capture the case detection effects through 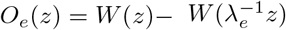 and 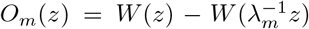. More general detection expressions require subtracting further time dilated terms involving the additional detection rates and demonstrate how detection probabilities and the generation time distribution are importantly coupled. The transfer function *G*(*z*) = *I*(*z*)*M* (*z*)^−1^ of (4) is a principal result that defines what a region acting in isolation can achieve using targeted interventions. We analyse ramifications of (4) in the Results.

### C. Interventions across source and sink locations

The framework developed above considers regions acting in isolation. Here we generalise (4) to factor in the infection dynamics within the source region. This allows us to model and assess coordinated interventions in the source and sink regions. Previously, our importations originated from the source region. We denote the mobility of infected individuals between source and sink regions using *ϵ*_*mi*_, *ϵ*_*im*_ ∈ (0, 1). At each time step, a proportion *ϵ*_*mi*_ of the newly infected individuals from the source migrate to the sink and a proportion *ϵ*_*im*_ of the newly infected from the sink migrate to the source. We note that *ϵ*_*ii*_ = 1 − *ϵ*_*im*_ and *ϵ*_*mm*_ = 1 − *ϵ*_*mi*_ are the proportions of new infections remaining in their original location.

The total number of newly infected individuals in the sink region at time *t* is then *i*(*t*) = *µ*_*mi*_(*t*) + *e*_*i*_(*t*), with infections resulting from contagion in the sink region, *e*_*i*_(*t*), satisfying the controlled renewal equation (5). Our notation differs slightly from (3) as we use *u*_*i*_ to indicate the proportion of net local *e*_*i*_(*t*) infections removed and *u*_*m*_ for the equivalent proportion of imported infections *µ*_*mi*_(*t*) = *ϵ*_*mi*_*e*_*m*_(*t*), which are removed. The cumulative detection probabilities are notated similarly and become 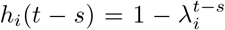 and 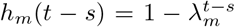 when detection is geometric. We use *R*_*i*_ = *ϵ*_*ii*_*R* and *R*_*m*_ = *ϵ*_*mm*_*R* to simplify formulae.

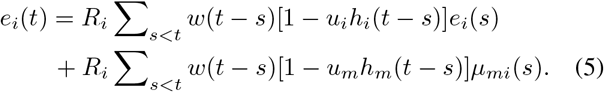

As we describe the dynamics of both source and sink regions, (5) is now accompanied by an additional controlled renewal model describing the net local infections generated at the source *e*_*m*_(*t*). This yields (6) with *µ*_*im*_(*t*) = *ϵ*_*im*_*e*_*i*_(*t*).

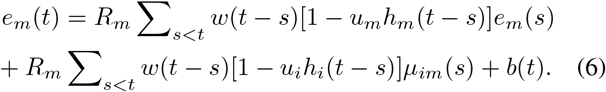

Because all source infections are subject to the same detection and control (i.e., imports in (5) are controlled in the source region), *e*_*m*_(*t*) faces detection *h*_*m*_(*t* − *s*) and removal proportion *u*_*m*_. The exports from the sink that enter the source region therefore also have been controlled via *u*_*i*_ and *h*_*i*_(*t* − *s*). We include *b*(*t*) as initial input infections to start the epidemic.

We take Z-transforms of our coupled renewal models to derive the matrix transfer function **G**(*z*) as in (7) where *I*(*z*) and *M* (*z*) are transforms of the total infections generated in the source and sink regions respectively.

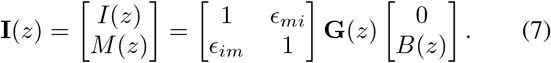

Here **G**(*z*) = (𝕀 − **Q**(*z*))^−1^, with 𝕀 as the identity matrix and **Q**(*z*) in (8). The diagonal terms of 𝕀 − **Q**(*z*) are similar to the denominator of our isolated region transfer function in (4).

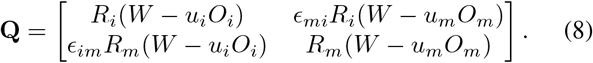

We examine the stability and performance of these equations in the Results to assess the impact of coordinated interventions.

## III. Results

### A. Border controls cannot stabilise epidemics

Equation (4) defines precisely how any importations drive the total number of new infections in our region of interest and immediately indicates why border closures or travel controls cannot stop epidemic growth. No import control terms feature in the denominator of (4), despite the *u*_*m*_ and *h*_*m*_(*t* − *s*) factors directly reducing ongoing and future transmission as in (3). The limited effect of controlling importations becomes obvious from the system block diagram of Fig. 1A. These interventions only alter the precompensator *K*_*m*_(*z*). Controllers *K*_*i*_(*z*) and *K*_*m*_(*z*) realise *E*(*z*)*M* (*z*)^−1^ with *I*(*z*) = *E*(*z*) + *M* (*z*).

**Fig. 1.**
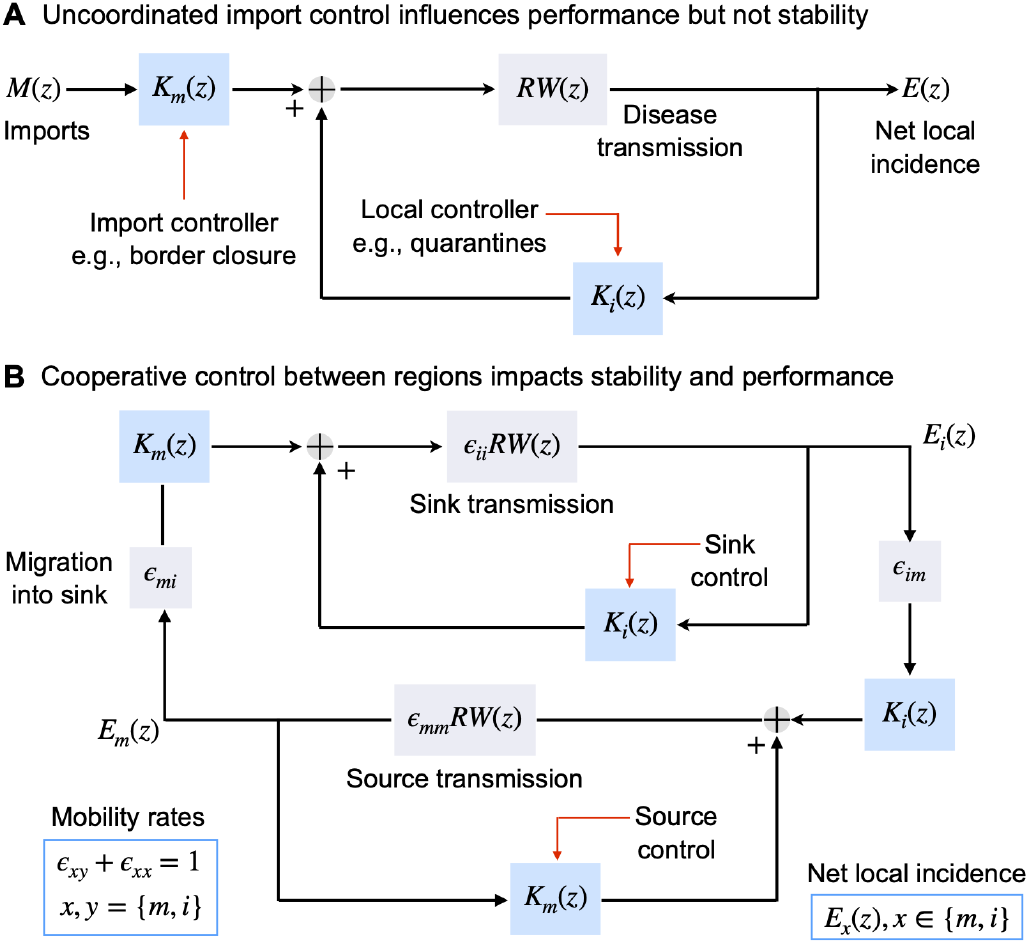
Control system architectures for managing imported and local infections. A: from the perspective of the local region with net local infections *E*(*z*), imported infections *M* (*z*) are disturbance inputs and border controls *K*_*m*_(*z*) always act as precompensators. This fundamentally limits the impact of *K*_*m*_(*z*) to shaping performance. Local measures *K*_*i*_(*z*) such as test-trace-isolate schemes always act in feedback with *E*(*z*) and define stability. Total infections *I*(*z*) = *E*(*z*) + *M* (*z*) and the controller forms are chosen to realise (4). B: under cooperative schemes *K*_*m*_(*z*) also becomes a feedback controller for the source region, which has total infections *M* (*z*) = *E*_*m*_(*z*) + *ϵ*_*im*_*E*_*i*_(*z*). This also implicitly adds an import control term for the sink region. Symmetrical constructions apply to the sink region with total infections *I*(*z*) = *E*_*i*_(*z*) + *ϵ*_*mi*_*E*_*m*_(*z*). The migration parameters mediate how infections are distributed. This structure with (*K*_*m*_(*z*), *K*_*i*_(*z*)) providing feedback control presents better (and likely more economical) solutions to suppressing the overall epidemic. We set the same *R* and *W* (*z*) for both regions for clarity, but this is easily generalised without issue. Controllers are chosen to realise (7) (but we do not show *B*(*z*)).

Moreover, as (4) is a linear system, provided that its poles are inside the unit circle or equally the controlled reproduction number *R*_*c*_ *<* 1, it also possesses bounded input-bounded output (BIBO) stability. As a result, if *m*(*t*) decays in finite time i.e., is transient, then *i*(*t*) decays regardless of the import control applied. Alternatively, if *m*(*t*) is growing, then import control cannot stop *i*(*t*) from growing, even at *R*_*c*_ ≪ 1. The only exceptions are unrealistic and trivial, requiring *R*_*c*_ = 0 or that all imports be removed upon arrival.

The poles of (4) satisfy *Ru*_*e*_*O*_*e*_(*z*) = *RW* (*z*) – 1. When *z* = − 1 solves this expression (as the dominant pole), the system is critically stable. Substituting this condition we find *R*_*c*_ = *R*(1 − *u*_*e*_*O*_*e*_(1)) and the new stability criterion of (9), linking detection and control to a herd immunity-type threshold *H*.

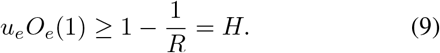

Hence, only the local controller *K*_*e*_(*z*) of Fig. 1A determines epidemic stability. The import controller has no influence on (9) but is important for shaping performance in the local region of interest as *K*_*m*_(*z*) controls the zeros of (4).

We define performance, given that *R*_*c*_ *<* 1, as the steadystate infection count *i*(∞) if *m*(*t*) is persistent, also known as the endemic equilibria, or the total new infections Σ_*t*_ *i*(*t*) if *m*(*t*) is transient (decaying), also termed the epidemic final size. Both quantities are of frequent public health importance. We compute the endemic state under a step input of size *m*(∞) by using the final value theorem. Setting a target ratio *ϕ* = *i*(∞)*m*(∞)^−1^, we derive the performance threshold (10).

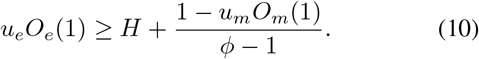

Equation (10) reveals that we must apply local control above that needed for stability from (9) by an amount based on our performance target (larger *ϕ* are easier to achieve) and our import control. We also find that (10) exactly describes final size performance with target *ϕ* = (Σ_*t*_ *i*(*t*))(Σ_*t*_ *m*(*t*))^−1^.

Interestingly, epidemic models also admit another endemic equilibria, which occurs at the critical *R*_*c*_ = 1 when driven by transient importations. This system features subexponential growth if imports are persistent. We apply the final value theorem with 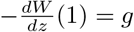 as the mean generation time to obtain 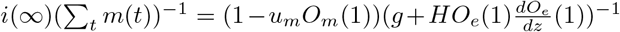. While not directly comparable to (10) as *u*_*e*_*O*_*e*_(1) is exactly *H*, we observe that import control is crucial to reducing the endemic infections via the factor 1 − *u*_*m*_*O*_*m*_(1) as in (10).

We explore and verify the above stability and performance thresholds for Covid-19, Ebola virus and Marburg in Fig. 2. Fig. 2A presents simulated epidemics with local control such that *R*_*c*_ *<* 1 and persistent step, sinusoidal and random import time series, all with the same steady-state mean. Import control substantially shapes performance, not only reducing endemic equilibria, but also damping peaks and oscillations. Fig. 2B plots how detection delays (mean *o*_*e*_ = *λ*_*e*_(1 − *λ*_*e*_)^−1^) can destabilise the control effort *u*_*e*_ required to achieve a specific *R*_*c*_ as in (9). Import control cannot shift these curves. Fig. 2C simulates epidemics at the critical *R*_*c*_ = 1 under decaying step, sinusoidal and random imports, validating how import control also shapes this second type of endemic state.

**Fig. 2.**
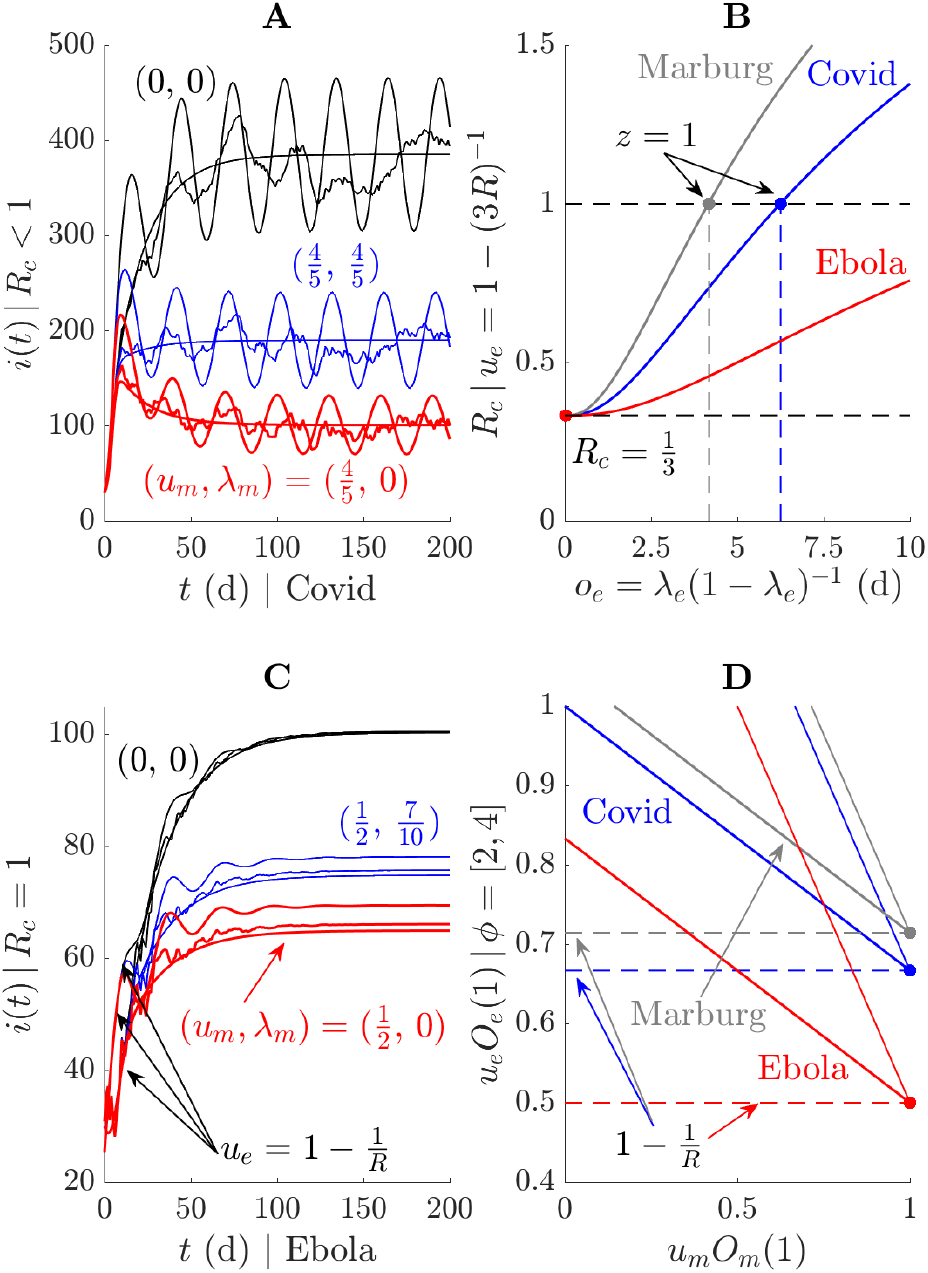
Performance and stability thresholds for uncoordinated strategies. A: epidemic trajectories *i*(*t*) simulated under Covid-19 parameters (from [21]) with stabilising local control (*R*_*c*_ *<* 1) and differing import control *u*_*m*_ and detection *λ*_*m*_ parameters. Persistent sinusoidal, step and random *m*(*t*) inputs are applied and responses are coloured by various import control settings. B: stability thresholds at local control *u*_*e*_ achieving *R*_*c*_ *<* 1 showing how geometric detection delays with mean *o*_*e*_ cause instability for various diseases (Ebola and Marburg parameters from [22]). These thresholds must be satisfied before focussing on performance as in A and C. Note that if the source region is unstable then these thresholds are not sufficient because imported infections are unbounded. C: epidemic trajectories simulated under *R*_*c*_ = 1 with Ebola parameters, coloured by different import control choices. Transient (decaying) sinusoidal, step and random *m*(*t*) inputs are applied and represent another way of attaining equilibria (as opposed to A). D: performance thresholds for achieving steady state and total infection ratios *ϕ* when *R*_*c*_ *<* 1 for various diseases. Import control i.e., *u*_*m*_ and *O*_*m*_(1) tuning, reduce how much local effort from *u*_*e*_ and *O*_*e*_(1) is needed above the stability threshold of B to achieve performance goals. Thinner lines are for the larger *ϕ* target, which requires smaller control effort. In all simulations of A and C, we run the uncontrolled model for a week before applying control.

In Fig. 2D we illustrate from (10) how import control can help reduce the local control effort needed to achieve a desired performance target *ϕ*. Perfect import control (*u*_*m*_*O*_*m*_(1) → 1) merges the control and performance criteria to *H* and makes explicit that a region acting in isolation, once stable, can limit importations to improve public health outcomes. This is an important point (in simulations *u*_*m*_ can even halve final sizes or endemic loads), suggesting that border closures and travel controls can be valuable tools for tuning performance.

Last, we show that the failure of border controls to stabilise epidemics is robust to important model variations. Specifically, (3) assumes importations acquire the transmission characteristics of the region they enter. This is a sink assumption. We may instead let importations maintain the transmission characteristics of the source i.e., reproduction number *R*_*m*_ and generation time distribution *w*_*m*_(*t*). Under this source assumption, the numerator of (4) becomes 1 +(*R*_*m*_*W*_*m*_(*z*) − *R*_*m*_*u*_*m*_*O*_*m*_(*z*)) − (*R*_*m*_*W*_*m*_(*z*) − *R*_*e*_*u*_*e*_*O*_*e*_(*z*)) but the denominator is unchanged. Consequently, even when the import region has *R*_*m*_ ≫ *R*, we cannot stabilise the epidemic with border controls. These points also hold for more complex source architectures (e.g., imports from multiple sources in series or parallel) because import control is always a precompensator for a BIBO system.

### B. Cooperative control provides stability and performance

As border and travel restrictions cannot stabilise infections, the only way to suppress epidemics is via cooperative source and sink control. The limitations of precompensation mean that when the source is unstable, there is no way for the sink region (acting alone) to stabilise infections. However, if we jointly consider sink and source regions, we can convert this precompensation effort into feedback control. This becomes apparent in Fig. 1B, which realises (5)-(6) and shows how net local infections in the sink *E*_*m*_(*z*) and source *E*_*i*_(*z*) are fed back to a suitable chosen *K*_*m*_(*z*) and *K*_*i*_(*z*).

The poles of the transfer matrix **G**(*z*) from (7)-(8) control the overall stability of the system (*i*(*t*), *m*(*t*)) and are the solutions to det (𝕀 − **Q**(*z*)) = 0. This leads to the characteristic polynomial Δ(*z*) of (11), generalising the denominator of (4).

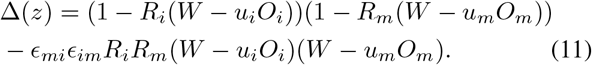

If either of the migration coefficients *ϵ*_*mi*_, *ϵ*_*im*_ equals 0, the poles are exactly the union of the poles of the isolated systems, and we recover the stability condition Δ(1) = 0 in (12).

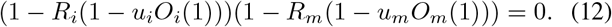

This produces the herd immunity type thresholds of *H*_*i*_ = 1 − (*ϵ*_*ii*_*R*)^−1^ and *H*_*m*_ = 1 − (*ϵ*_*mm*_*R*)^−1^, which are analogues of (9). If both migration terms equal 0, we recover *H*_*i*_ = *H*_*m*_ = *H*, where *H* is the threshold found in (9) in the absence of cooperative control. Importantly, when migration terms are non-zero, which is also necessary for *µ*_*mi*_ and *µ*_*im*_ to be realistic, coordinated interventions between regions can shift these thresholds and offer improved stability bounds.

We demonstrate this in Fig. 3A. When *u*_*i*_*O*_*i*_(1) ≥ *H* and *u*_*m*_*O*_*m*_(1) ≥ *H*, the uncoordinated locations are stable (Δ(1) = *ϵ*_*mi*_*ϵ*_*im*_(1 − *ϵ*_*mm*_*ϵ*_*ii*_) *>* 0). However, the coordinated domain of stability extends beyond these thresholds, being bounded by the hyperbola {Δ(1) = 0}, which has the lines {*u*_*i*_*O*_*i*_(1) = *H*_*i*_} and {*u*_*m*_*O*_*m*_(1) = *H*_*m*_} as asymptotes. As a result, for given levels of detection *λ*_*i*_ and *λ*_*m*_, cooperation between the source and sink region allows for smaller proportions of infections to be removed (and likely smaller intervention costs) while maintaining stability. In isolation, these control levels would result in unstable dynamics.

**Fig. 3.**
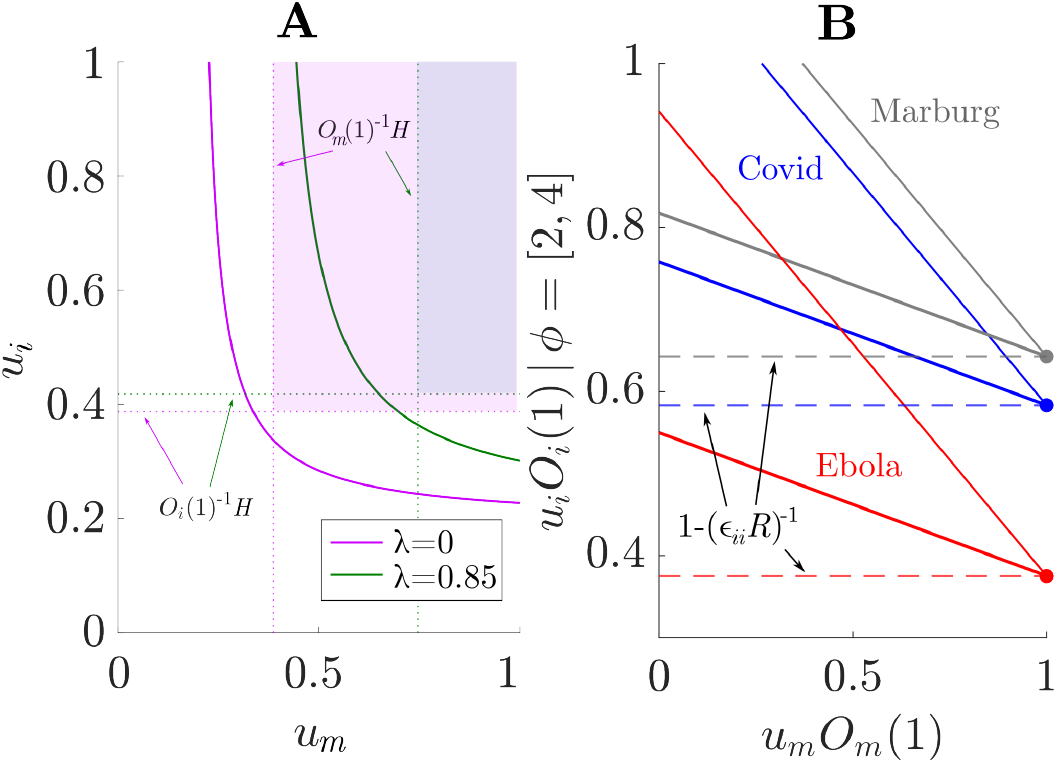
Performance and stability thresholds for coordinated strategies. A: Stability threshold curves {Δ(1) = 0} for *R* = 1.6, *ϵ*_*mi*_ = *ϵ*_*im*_ = 0.2, with *λ*_*i*_ = *λ*_*m*_*/*2 to illustrate a system with higher detection delays in the sink region. The asymptotes of the stability curves are the herd immunity thresholds *H*_*i*_ and *H*_*m*_ for the isolated systems when one of the *ϵ*_*mi*_, *ϵ*_*im*_ equals 0. Shaded regions represent the stability regions for the system without cooperative control given by (4). Gaps between a curve and its corresponding shaded region indicate the benefits of cooperative control. B: performance thresholds for achieving steady-state and total infection ratios *ϕ* for various diseases, as in Fig. 2. Stability regions (as shown in panel A) and threshold curves are much lower than in the equivalent uncooperative system, indicating the feasibility of effective performance management with lower control effort in the case of cooperative control (provided stability is achieved).

This improvement becomes more appreciable (i.e., the asymptotes are farther from the isolated thresholds) as detection delays become larger. Moreover, as 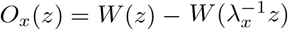 for *x* ∈ {*i, m*}, is increasing in *λ*_*x*_, (11) reveals that for a specific mobility structure *ϵ*_*im*_, *ϵ*_*mi*_ and fixed transmission parameter *R >* 0, outbreaks can also be stabilized by decreasing the delays in detection, even at fixed control levels *u*_*i*_ and *u*_*m*_. The benefits of coordination on stability are stark.

We can also assess performance, from the perspective of the sink region, by considering the steady-state infection count *i*(∞) under a step input *b*(*t*). This can be computed using the final value theorem to derive (13).

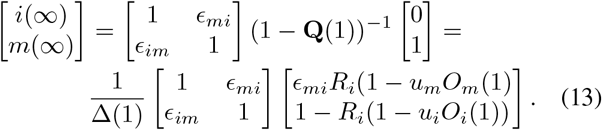

We set a target *ϕ >* 1 for the ratio of infections in the sink region to the number of incoming infections from the source region, in line with (10). Considering only those infections that remain in the sink region and that only a fraction *ϵ*_*mi*_ of the total steady-state infections in the source region migrate to the sink, this means that we need to set controls such that (*ϵ*_*ii*_*i*(∞))*/*(*ϵ*_*mi*_*m*(∞)) = *ϕ*. Given the explicit solution from (13), we represent the necessary control levels in Fig. 3B for contagion levels associated to different diseases.

Again, in the limit of perfect control in the source region (*u*_*m*_*O*_*m*_(1) → 1), for all target ratios *ϕ*, the necessary level of control in the sink region converges to the minimal level *H*_*i*_ = 1 − (*ϵ*_*ii*_*R*)^−1^ that still guarantees stability. Remarkably, we find that the performance curves in Fig. 3B are uniformly lower than their equivalent in Fig. 2D (within the bounds of stability). This conclusively illustrates that cooperative control in interconnected regions not only improves global stability of the system, but also achieves similar levels of performance at lower control efforts. These benefits are important given the high financial and societal costs of epidemic control measures.

## IV. Discussion

We have demonstrated conclusively that border controls and travel restrictions cannot stabilise epidemics, simply because they do not alter the dominant pole of the epidemic. Regions acting in isolation can only meaningfully reduce spread by implementing local interventions that remove a proportion of infections above a herd immunity-type threshold *H*. However, even these measures fail if the importations originate from an unstable region (due to limits imposed by BIBO stability). Consequently, overall stability is best achieved by coordinating control strategies among interacting regions.

This supports more complex models from [8] suggesting that countries should share resources (e.g., vaccine stockpiles) and cooperatively respond to epidemics. We found that such joint control actions reduce the overall control effort required for stability and performance. Cooperative strategies notably expand the parameter space for stability (Fig. 3A), meaning that lower levels of control effort in both source and sink regions can collectively stabilise the system, even under conditions where the isolated regions would remain unstable.

Although border closures fail to suppress outbreaks, they can be valuable, once stability is ensured. The aim then becomes to minimise total and endemic infection loads. Achieving such performance usually requires local interventions of strength substantially above *H*. Border controls can make this problem considerably easier and even restore the lower bound of *H* (Fig. 2D). These benefits are realised both by isolated and cooperative strategies, but achieving performance targets demands less stringent measures in each location when acting jointly compared to isolated action (Fig. 3B). Similar phenomena have already been discovered in models of networked agents [23], but had not yet been made explicit using control theory applied to generalised epidemic systems.

Several simplifying assumptions underlie these results. First, we neglected all stochasticity. During early epidemic stages, imported infections can randomly fade out (failing to trigger epidemics) or generate unexpectedly large outbreaks (superspreading) [24]. While these are key aspects of importation dynamics, they likely only set the timing of epidemic waves. Once an epidemic takes off we expect our analyses to be a fair approximation of dynamics. Second, we do not consider the non-linear effect of susceptible depletion. However, this is mostly an issue around the epidemic peak and model linearisation is common for initial and late epidemic stages. Last, we do not model the additional and hard-to-quantify feedback loops between control actions and population behaviours. This is a non-trivial effect (e.g., behaviour can alter import patterns [25]) that we aim to incorporate in future renewal models.

More broadly, the interaction between migration and heterogeneous control policies that are applied across different locations is a complex issue, which we have viewed through the simplifying yet insightful lens of two interacting locations. For larger systems with more complicated migration matrices, identifying the combinations of local interventions and border controls that achieve stability and performance goals becomes even more challenging [26]. Devising optimal cooperative strategies in such settings remains an open question that we aim to explore further using the foundations laid by our controlled renewal approach, with the goal of providing guidance for pandemic management across interconnected regions.

## Data Availability

There is no data associated with this article.

## Acknowledgment

KVP thanks Richard Pates for helpful discussions on modelling and controlling epidemics.

